# Multivariate Prediction Network Model for epidemic progression to study the effects of lockdown time and coverage on a closed community on theoretical and real scenarios of COVID-19

**DOI:** 10.1101/2020.05.04.20090712

**Authors:** Dimitri Marques Abramov, Saint-Clair Gomes Junior

## Abstract

The aim of this study was to develop a realistic network model to predict the relationship between lockdown duration and coverage in controlling the progression of the incidence curve of an epidemic with the characteristics of COVID-19 in two scenarios (1) a closed and non-immune population, and (2) a real scenario from State of Rio de Janeiro from May 6^th^ 2020.

Effects of lockdown time and rate on the progression of an epidemic incidence curve in a virtual population of 10 thousand subjects. Predictor variables were reproductive values established in the most recent literature (R0 =2.7 and 5.7, and Re = 1.28 from Rio de Janeiro State at May 6^th^), without lockdown and with coverages of 25%, 50%, and 90% for 21, 35, 70, and 140 days in up to 13 different scenarios for each R0/Re, where individuals remained infected and transmitters for 14 days. We estimated model validity in theoretical and real scenarios respectively by applying an exponential model on the incidence curve with no lockdown with growth rate coefficient observed in realistic scenarios, and (2) fitting real data series from RJ upon simulated data, respectively.

For R0=5.7, the flattening of the curve occurs only with long lockdown periods (70 and 140 days) with a 90% coverage. For R0=2.7, coverages of 25 and 50% also result in curve flattening and reduction of total cases, provided they occur for a long period (70 days or more). For realistic scenario in Rio de Janeiro, lockdowns +25% or more from May 6^th^ during 140 days showed expressive flattening and number of COVID cases two to five times lower. If a more intense coverage lockdown (about +25 to +50% as much as the current one) will be implemented until June 6^th^ during at least 70 days, it is still possible reduce nearly 40-50% the impact of pandemy in state of Rio de Janeiro.

These data corroborate the importance of lockdown duration regardless of virus transmission and sometimes of intensity of coverage, either in realistic or theoretical scenarios of COVID-10 epidemics. Even later, the improvement of lockdown coverage can be effective to minimize the impact of epidemic.

## INTRODUCTION

SARS-CoV2 epidemic has had a significant impact on global public healthcare, due to its high transmissibility and to the significant mortality of COVID-19 (Rothan and Byrareddy, 2020). The pandemic was declared on March 2020 by the WHO (Cucinotta and Vanelli, 2020), mobilizing all nations to take lockdown measures, since there is still no effective treatment for this pandemic.

Economic and social costs of this lockdown must be considered in the projection of which type of lockdown must be adopted. Observing the progression of the pandemic in its epicenter, in March, Shen & Bar-Yam, from the New England Complex Systems Institute (2020), estimated that an extreme lockdown of 5 weeks was sufficient to contain the COVID-19 epidemic in China, warning that a soft lockdown would be ineffective. The ideal lockdown coverage curbs new infections while it buys time for the virus to die out in individuals who are already infected, (1) significantly reducing the infected population, and (2) producing immunity barriers, thus interrupting infection spread (Kissler et al, 2020). These predictions have been based on mathematical modeling using epidemic characteristics, such as initial reproductive number (R0) and effective reproductive number (Re) (Kissler et al., 2020; Zhao and Chen), or epidemic growth rate (Wibens et al, 2020).

As opposed to models based on linear equations, Network Models are adequate for realistic simulations of complex systems with dynamic behavior patterns, where the individual states of its units and the connections among them are intrinsically non-linear, and emulated by pseudo-random series (Haykin, 1994; Britton, 2020).

Even though it is nearly impossible to determine objectively the ideal, or even realistic, parameters for a model (regarding R0 and quantification of effective lockdown coverage), we might still estimate the relationship between lockdown time and coverage in truthful scenarios. Moreover, these predictions would be important at least as motivation for the planning of public lockdown policies. Coverage is an objectifiable measure in terms of proportions (in %) for comparisons between scenarios with shared parameters, which has qualitative value. In the present evaluation, the most important parameter in this simulation is virus reproduction rate in a non-immune population (R0). R0 for COVID-19 had been estimated as 2.7 (Wu et al, 2020). However, new estimates on pandemic progression have obtained a R0 of 5.7 and a growth rate of 0.21-0.3/day (Sanche et al, 2020). Other recent sources have indicated reproductive numbers of this magnitude (Bulut and Kato, 2020; Zhao and Chen, 2020). In Rio de Janeiro, the current effective reproductive number is estimated about 1.44 (Melan et al., 2020).

Therefore, we developed a network model to estimate the effect of lockdown coverage intensity and coverage time on incidence curves of COVID-19 epidemic in a closed population (with no significant exchange of individuals from/to other populations). Our intention is to check how these two dimensions might comparatively affect amplitude and latency of peak incidence, as well as total number of cases in theoretical scenarios. For that, we adopted a small population of 10 thousand non-immune individuals in the simulations, observing incidence progression over 1000 days, considering the conservative and pessimistic R0 estimates of 2.7 and 5.7 transmissions, respectively, per infected individual, on average, as central parameters

We performed simulation for the State of Rio de Janeiro in this virtual population, that is a realistic scenario. We aim to predict the effects of different lockdown coverages and time periods. To do it, we use the most current data about the number of infected people and the effective reproductive number estimated on May 6^th^, by Mellan et al. (2020).

## METHODOLOGY

We conducted a theoretical study on the progression of the incidence curve comparing possible lockdown coverage and time scenarios, concerning peak amplitude, peak latency, and total number of cases. Considering the two R0 values estimated in literature for COVID-19 (Sanche et al, 2020), the study scenarios are as follows: no lockdown (natural progression of the epidemic); and lockdowns with proportional coverages of 25, 50, and 90% in periods of 21, 35, 70, and 140 days, with a total of 13 scenarios for each R0 value. Realistic simulations had used parameters obtainded from Mellan et al (2020).

Our period of analysis was arbitrarily defined as 1000 days, with temporal accuracy of 1 day. We applied lockdown scenarios starting from the first day of simulation, with the specified coverages and periods.

We used a Network model to simulate the dynamics (progression) of changes in the state (non-immune, infected, and immune/deceased) of the units (population subjects) through their mutual connections, by which the infection spreads.

The universe of this model is a virtual, random-sized population with 10 thousand subjects socially interconnected. We interpreted these connections as the likelihood of each subject infecting each one of their peers. The number of infected subjects per each subject might vary from 0 to infinite. These connections are stochastically based. Thus, due to the non-linear nature of the network model, we used a sample with thirty simulations of each of the 26 scenarios studied, starting from different “patients zero”. As criterion for patient-zero eligibility, we adopted patients who had a likelihood of transmitting the disease different from zero.

The script developed in Octave/MatLab language is found as supplementary information for free use, in the repository: https://data.mendeley.com/datasets

### Model structure

We adopted a vector **u** with 10 thousand values (N = 10000), representing a population with 10 thousand subjects *i* who were non-immune (u(i) = 0), many of whom shall be infected (u(i)=1), progressing to the outcome (u(i) = NaN, *not-a-number*), once they become immune or die. Epidemic progression is determined by the total amount of individuals *i* who are infected and progress to the outcome in each time unit *t* (in hypothetical days), with *t* = 1,…,1000.

The N individuals *i* established connections *Cij* with individuals *j* in a non-bidirectional manner (*Cij ≠ Cji and Cii = NaN*). Connection matrix **C** determines the likelihood of a subject *i* infecting other subjects *j* (*j = 1,…, N*-1), according to a probability density function (pdf) *T* with *N*-2 degrees of freedom (figure 1). The number of subjects *j* who might be infected by the individual *i* is determined by a pseudo-random series **r** with size *N*-1, and lambda Poisson distribution equal to *R0*, according to pdf T. In other words, the infection between *i* and *j* shall occur according to the conditional function:

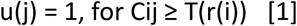

**Figure 01.**
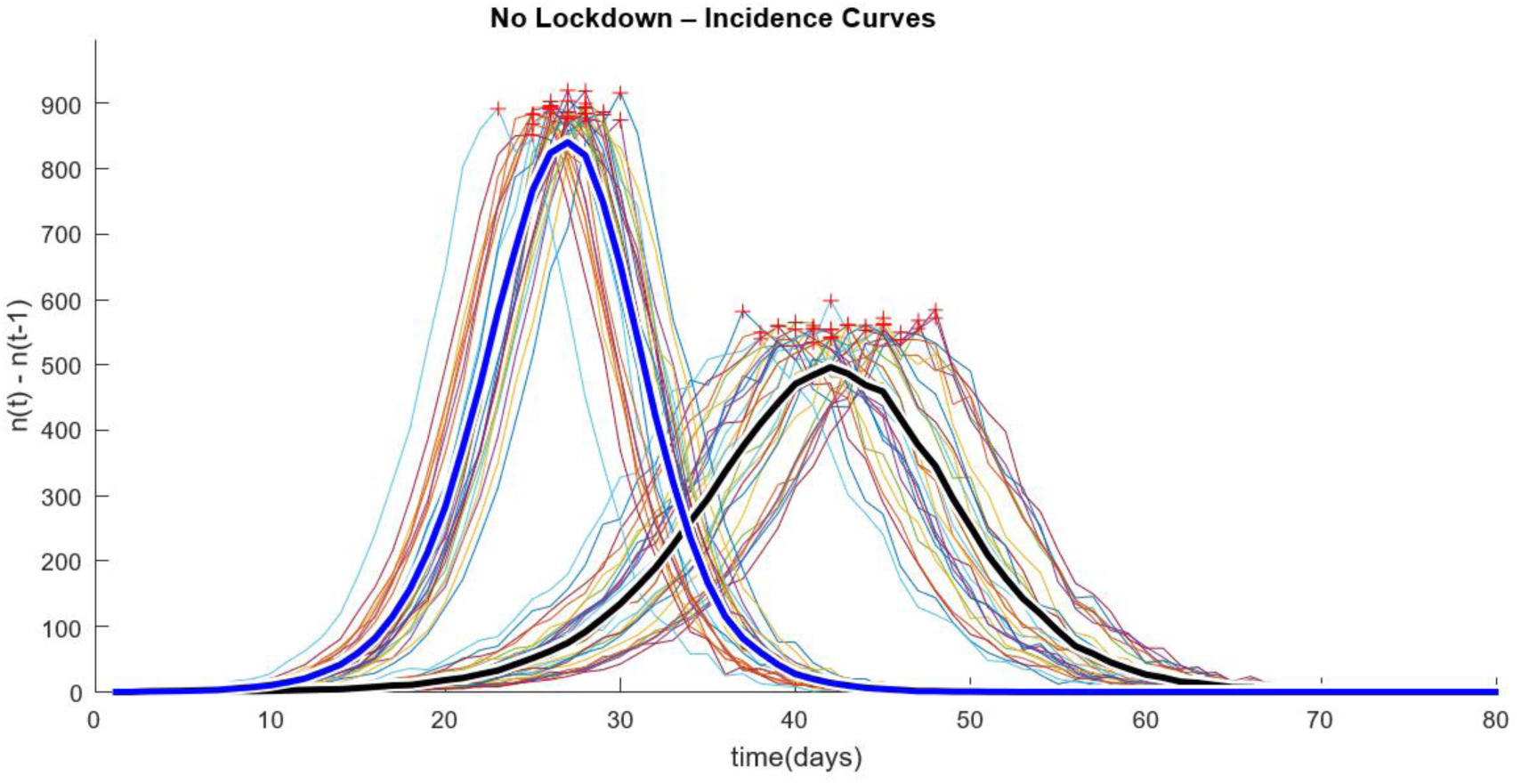
Incidence curves in a sample with thirty simulations for R0 of 2.7 and 5.7. Mean values are plotted above the corresponding samples, in black and blue, respectively. Peaks are indicated with red crosses. See table 01.

The weight of connections *Cij* had a random uniform distribution, leading to a topographic connectivity spread.

### Model parameters - predictors

The parameters and values adopted in this model were:

*Virus reproduction number in a non-immune population:* **R0** = [2.7, 5.7], according to Wu et al. (2020) and Sanche et al (2020), respectively. This parameter is dynamic in practice, as the non-immune population decreases over time, becoming *Re* (effective reproductive number) (Aronson et al, 2020). For simulations for Rio de Janeiro, according Mellan et al. (2020), on May 6^th^ the effective reproductive number (Re) in Rio de Janeiro State (RJ) was 1.44 (C.I. 1.28, 1.60). We found the **Re** inside this interval that yields the epidemic dynamics which best fit to real data realizing simulations without lockdown (procedure and data not shown).

*Days of individual infection progression: p* = 14, which is the mean period of infection and transmission by COVID-19 and we presumed that 50% of infections occur between the fourth and eighth days, when the elimination of viral particles is higher (Cevik et al, 2020).

Finally, the parameters analyzed are *lockdown period*, **δ** = [21, 35, 70, 140], in days, and *relative lockdown coverage*, **α** = [Inf, 4, 2, 1.12], corresponding to no coverage, 25%, 50%, and 90% of coverage, respectively.

In the model dynamics, lockdown is the reduction in values of matrix **C** proportional to α(k) during a period δ(k), where *k* = 1,..,4 and *t* = 1,…, δ, i.e., lockdown always on the first day of epidemic progression in population u, which is characterized by the confirmed first case of community transmission, as described in the next section. Values **C** are determined according to the conditional function below:

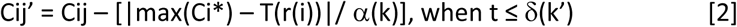

Where *k* and *k’* progress independently.

For realistic scenario in RJ, we also realized a simulation for lockdowns from June, 6^th^ onwards.

### Model dynamics

First, one individual *i* is randomly selected to be the first patient with community transmission, *u(i)* = 1 at *t*=1.

In simulations for Rio de Janeiro, we randomly selected a proportional number of initially infected (u(i) =1) and immune/deceased subjects (u(i) = NaN) as initial conditions (t=1). Estimating the number of total cases about 582000 (C.I. 492000, 657000) with 1285 deaths. The recovery rate to Rio de Janeiro is about 70% (Sec Estado Saúde RJ, 2020), what is, about 173000 people is infected now and 409000 is immune or deceased. The state population was estimated in 17.264.943 (IBGE, 2019). Thus, about 3,4% of all state people could be infected by SARS-CoV2, where roughly 1,0% remain active. We feed our model with this mean Re (1.28) and we randomly turned 240 subjects as immune/death (not-a-number) and 100 people are active at t=1, as estimated for May 6^th^ (.

From this point on, each infected subject remains infected and transmitting over the period *p*. A vector, **π**(1: p) = 0, is initially determined, representing *π* days of infection. Infections are distributed semi-randomly in **π**, according to the following conditional function:

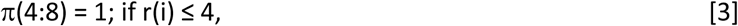

*π*(4:8) = 1 and *π*(rnd^p^(r(i)-4)) = *π*(rnd^p^(r(i)-4)) +1; otherwise

Where *rnd* is the generation function of r(i)-4 random values between 1 and *p*. In other words, it is possible to have *1:r(i)-3* infections on one single day. In the course of time, *t*, subjects *j* are infected by *i* while 1 < *Δt* < *p*, in a decreasing likelihood order (first, the most likely), given pdf *T*. Each u(j) = 1 starts a cycle of *p* days of transmission, until *u(j)* = NaN, if *Δt - p* ≥ 1.

In each cycle (*Δt* = 1), subject *i* infected new subjects *j* at *(t-1, t-13)*. Each infected subject *i* becomes immune/deceased with *Δt* = 14, turning into *u(i) =* NaN. At the end of the iteration by all *N* subjects, the current status **u** is stored.

### Outputs and Analysis

As outputs for analysis, we evaluated the total number of infections per day, *n(t)*, where *t = 1,…,1000*, from which the curves of new infections *(n(t) - n(t-1))*, called incidence curves, and the curve of epidemic growth rate *(n(t)/n(t-1))* are derived.

In order to validate the model in theoretical scenario, we calculated coefficients of Growth Rate, *r_Ro(1_*_)_ and *r_Ro(2)_*, through the best-fit curve of an exponential model (Ma et al., 2013), applied to the incidence curve:

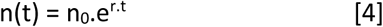

Where *n*_0_ = 1 (initial number of infected subjects), t is the time in days, and *r* is the exponential growth coefficient of the curve. In order to apply the best-fit model, we varied the *r*-value from 0.05 to 0.35 with 0.005 increases.

Given the pseudo-random nature of the network model, we performed 30 to 50 simulations by theoretical and real scenarios respectively, raffling the corresponding initial patients, for each parameter, **R0**, **δ** and **α**, which determined up to 26 groups, since parameter **δ** is irrelevant for *a(1) = Inf* (in lockdown).

The values calculated were peak amplitude of the curve of new cases, peak latency, and total number of infected and outcomes in **u**, when *t* = 1000. Peak latency of each output “k” was weighed to correct distortion effect in the sample, and it was multiplied by the corresponding peak amplitude and divided by the mean amplitudes calculated in the output sample.

In order to describe the magnitude of the inherent non-linearity of the model, we showed sample statistics in terms of median and percentis (5% and 95%) for theoretical scenarios, discriminated according to the R0 adopted (table 1).

**Table 01.**
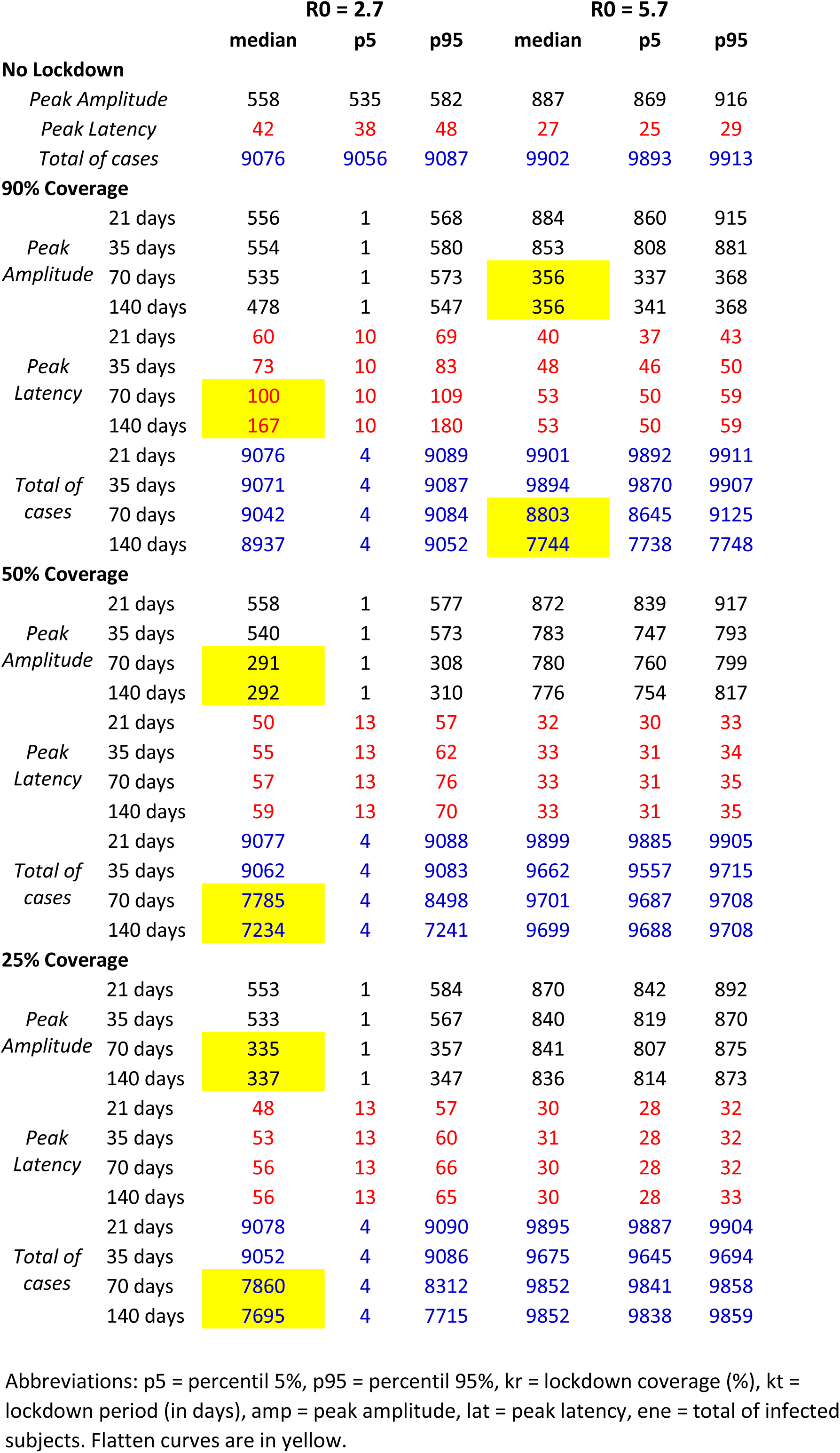
descriptive statistics regarding R0 values groups (n=30): peak amplitude and latency of incidence curve (n(t)-n(t-1)) and total of cases (total prevalence).

Graphs are the mean values plus two standard deviation of each realistic scenario. Due to the accuracy and predictability inherent to models, inferences are irrelevant since likelihood of equality shall always be inversely proportional to the size of samples of simulations.

Regarding simulations for real scenarios, despite our small population size, the model is representative if the behavior of real population is homogenous and its dynamics is isotropic, as in the model. That is, if populational mobility from/to outside the state is not relevant. For estimates, we must re-scaling model data to real data dimension.

## COMMENTED RESULTS

### PART I: POPULATION DESCRIPTIONS

Using pseudo-random Poisson distributions (see methodology) to adjust the likelihood of transmission in the connections between subjects, we found that the maximum number of subjects infected by the same subject was 7, 11 and 17 for Re=1.28, and R0 = 2.7 and 5.7, respectively. Additionally, the proportion of non-transmitters was 23.84%, 6.9% and 0.23% for these reproductive number, respectively.

### PART II: SIMULATIONS ON THEORETICAL SCENARIOS TO COVID-19 EPIDEMY

Ideal and homogeneous and isotropic population of 10 thousand non-imune individuals in 30 simulations whose initial conditions are one patient-zero randomnly chosen per simulation, using R0=2.7 and 5.7. We collectively consolidated individual outcomes (having been infected or not) by the end of the 1000 simulated days for a descriptive analysis of each lockdown scenario, and the comparison between them, as shown in table 1, where we described data in terms of median and percentis 5% and 95%.

#### Validating the model using growth rate coefficient

The reliability of the model is indicated by the comparison between the growth Rate (*r*) estimated by Sanche et al (2020) for *R0* = 5.7 and the growth rate observed. The simulation shows dynamic rates, which is perfectly predictable considering that the effective variation of R0 into Re was progressively lower than *R0*, until epidemic progression was stabilized, when *n(t)-n(t-1)* = 0. When *R0* = 2.7, the epidemic progresses more slowly, maintaining an *r* lower than that for *R0* = 5.7.

Using the exponential model (eq. 4), we found *r(R0=2.7)* = 0.16 and *r(R0=5.7)* = 0.28, according to the incidence curves, i.e. the progression of new infections and individual outcomes. The most recent *r* estimates occurred in the interval 0.21-0.3/day, related to *R0=* 5.7, as opposed to the previously estimated r, of 0.1–0.14, related to *R0* = 2.7 (Sanche et al, 2020).

#### Behavior of epidemics without lockdown

As the incidence curves in figure 01 show, the epidemic progresses more rapidly with R0 = 5.7, establishing a higher and earlier peak of infection than with R0 = 2.7. Both infections result in more than 90% of subjects contaminated until immunological barriers are established.

#### Behavior of incidence curves during Lockdown

Descriptive data of peak amplitude, peak latency, and total number of infected subjects (medians and percentis 5% and 95%) for outputs related to R0 equal to 2.7 and 5.7 might be seen in table 01. The first observation is related to the wide data dispersion when the epidemic with R0=2.7 has a 90% coverage, because the likelihood of an epidemic not progressing increases as patient zero might have very thin connections with other subjects.

##### Effect of lockdown coverage

Considering R0 = 5.7, intensity of coverage (25%, 50%, or 90%) had a visible effect on curve peak amplitude and latency during lockdown of 70 days or 140 days, although it was relevant only for coverages of 90%, when the curve was effectively flattened (see bellow). Coverage intensity also affected the total number of infected subjects, although a relevant reduction, of approximately 12% of the infection cases (see table 01), only occurred with 90% coverage.

When we consider results for R0=2.7, we observe that any coverage intensity affected curve behavior compared to no lockdown. For 25 and 50% of coverage, there was flattening of the curve too (peak amplitude 40% to 50% lower than no lockdown) with observed effect of approximately 20% on the total number of cases compared to no lockdown (see table 01).

##### Effect of lockdown duration with 90% coverage

We evaluated the effect of lockdown period (21, 35, 70, or 140 days) on the incidence curve, considering the ideal lockdown (90%). The data are described on Table 1. We observed that all curves are different from each other in peak latency and amplitude for R0=5.7, except for peak amplitude related to the period of 21 days compared with no lockdown. The peak latencies related to 70 and 140 days of lockdown overlap, as they are the only ones that showed a relevant flattening effect on the incidence curve (amplitude almost 60% lower compared with no lockdown). Considering R0=5.7, a lockdown of 21 and 35 days with 90% of coverage only delays incidence curve peak by a few days, and longer periods of time are required to flatten the curve.

For R0 = 2.7, lockdown with 90% of coverage was only effective in delaying the peak latency, more significantly with 70 and 140 days of lockdown, although there was no flattening of the curve (peak amplitude only 1% to 15% comparing with no lockdown). Periods of 21 and 35 days, once again, seem to have been effective to delay the incidence curve peak. Lockdowns of 70 and 140 days reduced the number of infected subjects only for R0 = 5.7.

##### Effect of soft lockdown with short duration

Coverages of 25 and 50% during 21 and 35 days did not have any relevant effect on the dynamics of epidemic progression, whether in terms of incidence curve peak amplitude and latency, for either R0=2.7 or 5.7, or in terms of total number of cases (table 1).

### PART III: SIMULATIONS ON REAL SCENARIO OF RIO DE JANEIRO AT MAY 6^TH^ 2020

#### Model Validation

As mortality is an index that suffers lower under-reporting, and (2) mortality/total cases is a fixed rate before health system collapse (thus indicating proportional progression of epidemy), we used cumulative mortality reports between May 6^th^ and 27^th^ (from Ministério da Saúde do Brasil, 2020) as indicator to validate the model (as made by Melan et al, 2020). The cumulative mortality was adjusted to best fit it upon predicted curve (red curve in figure 2):

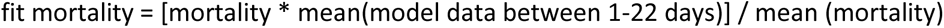

**Figure 2.**
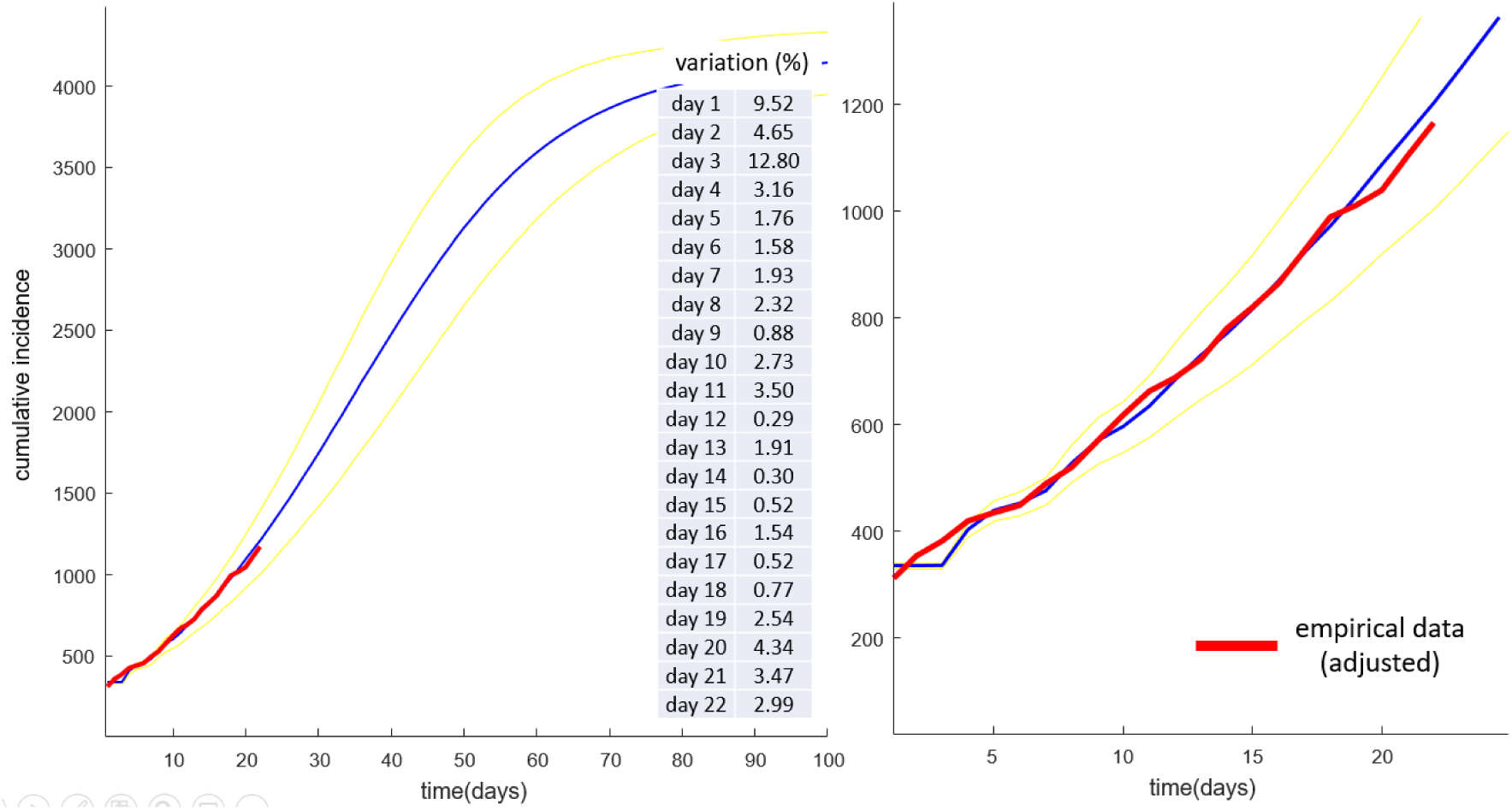
Evidence of validity – real data series upon modeled data from may 6^th^ to 27^th^ for State of Rio de Janeiro. Red: adjusted data about cumulative mortality (see text). Blue: predicted data about epidemy progression (cumulative cases, mean of 50 simulations). Yellow: ± 2 standard deviation of mean. Table: variation rate (%) between real (normalized) and predicted data.

By simulations without lockdown (results not presented), the best reproductive number found was 1.28, inside confidence interval for Rio de Janeiro (Mellan et al, 2020). The variation between empirical and simulated data was from 0.29% to 12.80%, with average equal to 2.97 % ± 3.25% (mean ± st.dev). See table in figure 2.

### Estimates for Rio de Janeiro, Brazil with diferent lockdown coverages

Under current status in Rio de Janeiro (effective reproductive number = 1.28, with approximately 3.4% of population already infected, 2.4% recovered/deceased and 1.0% with active disease), the epidemic progresses rapidly reaching its peak in 32.66 + 3.32 days (mean+std. dev). Total infected subjects will be 54.03% ± 0.05% of State population (nearly 8.5 million of people) roughly in next 90 days from May 6^th^. See figure 03. Regarding mortality from May 6^th^ (1205 deceased people), we can estimate a total of 14871 ± 588 deaths (mean ± standard deviation) directly caused by COVID-19.

**Figure 3.**
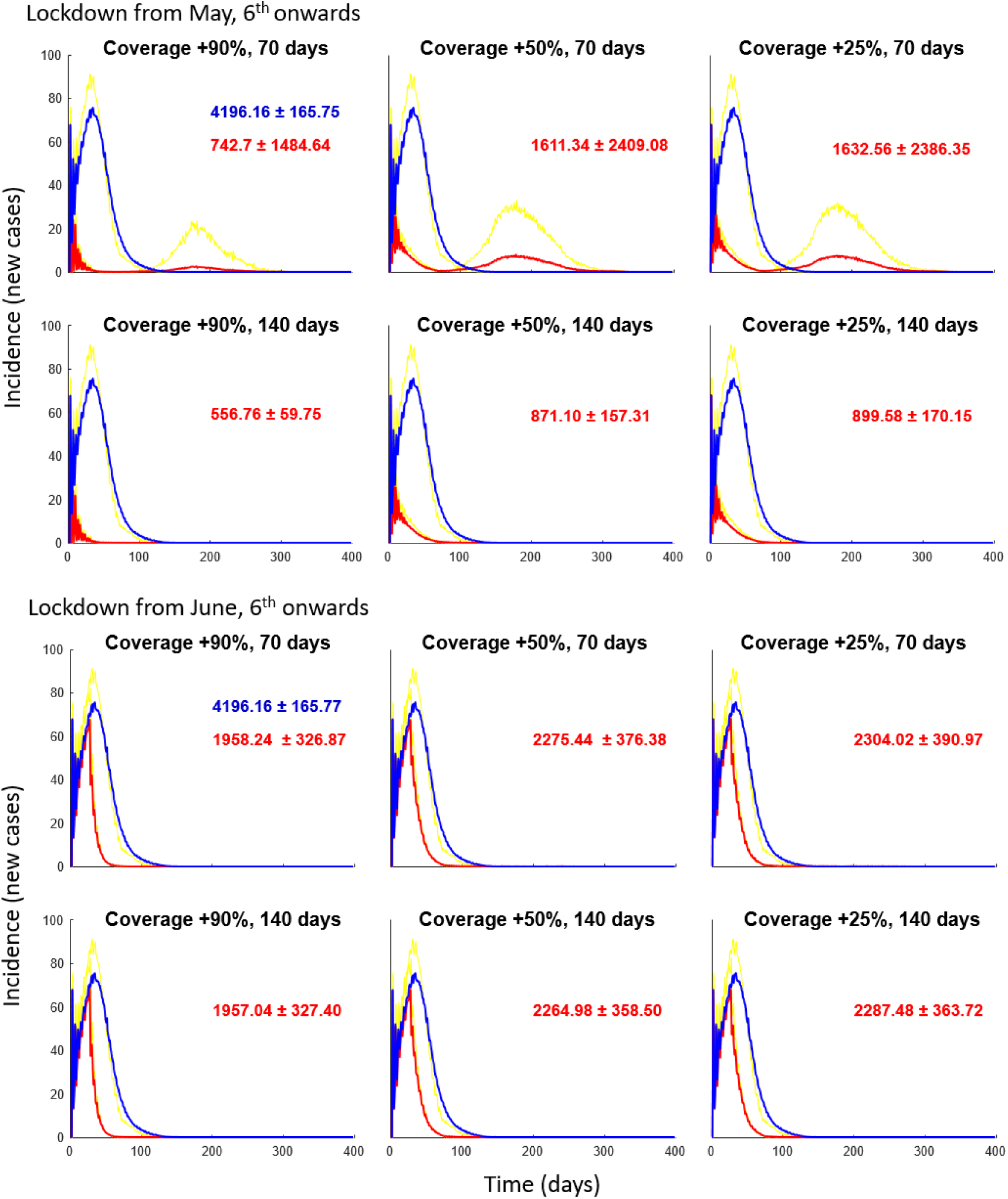
Effect of Lockdown Coverage(+%) and duration with Re = 1.28 (Rio de Janeiro scenario). Initial conditions: 100 active and 240 imunne/deceased (see text). 50 simulations. Blue: No lockdown, red: respective lockdown parameters (subplot title). Yellow: standard error of mean. In plots, total of cases (mean ± 2 std. dev).

**Figure 4.**
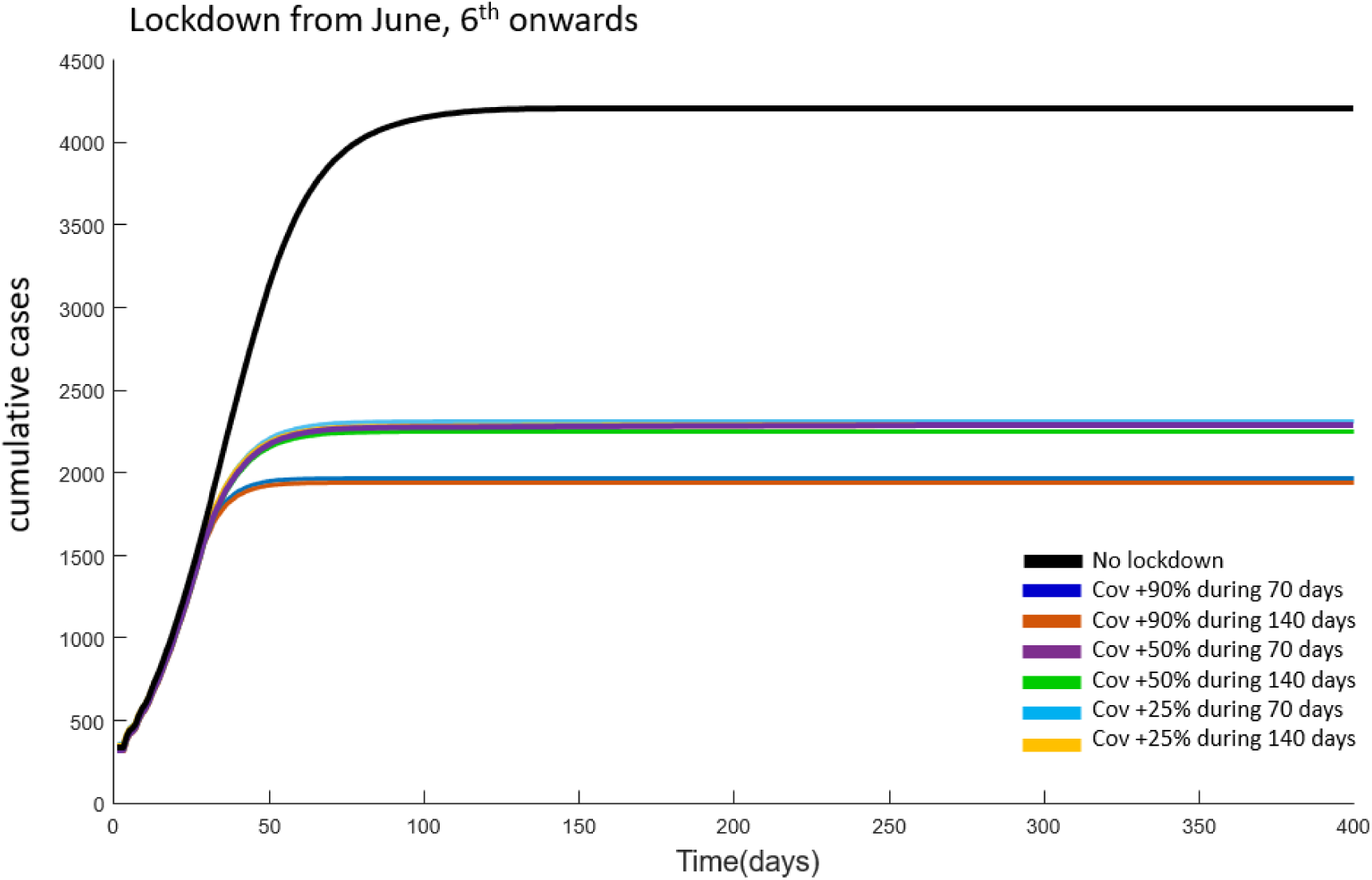
Effect of Lockdown Coverage(+%) and duration beginning at June 6^th^ at Rio de Janeiro State, with Re = 1.28: cumulative cases. Initial conditions: 100 active and 240 imunne/deceased (see text). 50 simulations. See color captions.

If increasing social confinement at first day (May 6^th^), the incidence softly would fall near to zero. However sooner or later the incidence would increase again, depending on the duration of the lockdown. With at least +25% of lockdown coverage during 140 days the incidence would modestly increase after lockdown, and the total number of cases will drop at least to 21.61% + 12.17% of State population. Coverages with duration of 70 days would flatten the curve but reduce the total number of cases nearly 10%. See figure 06. In line with results above described, the main determinant of lockdown efficacy would be its duration, since coverages of plus 25 or 50% show similar effects. Really, +90% of coverage (practically a total lockdown) would drastically reduce total number of infected subjects.

Predictions about an additional lockdown from June 6^th^ reveal effectivity against the epidemic. With plus 25% coverage (near 65 to 70% of lockdown) at next June 6^th^, this number falls to nearly 8000 deaths (against 14871 ± 588 without any new measure). Proportionally, we can infer about total of cases: without additional coverage, nearly 41% of population will be infected by SARS-CoV2. This prevalence would fall nearly from 22 to 23% of total state population. In this case, lockdown duration (70 or 140 days) has no relevant effect.

## DISCUSSION

Network models are frequently used in simulation of neural and cognitive processes, and are a quite realistic modelling strategy for the dynamics of a complex collectivity, as they simulate dynamic information and energy flows through the connection matrix between Network units (Haykin, 1994). With adequate parameters, a network model virtually behaves as a real life population.

In order to simulate realistic scenarios, which have an intrinsic non-linear nature, we used here generators of stochastic numbers: raffles shall always be required to define who is connected to whom and with what intensity. In this application, elucidating behavior patterns of the simulated collectivity occurs by empirical observation of the progression of this interconnected collectivity. In this case, the non-linearity is in the connection matrices (which are here interpreted as the likelihood of subjects being contaminated). Therefore, we worked with output samples, derived from different simulations: several evolutions are possible as initial conditions change. So, we must work with a larger sample of outputs as possible from different simulations, varying initial conditions (i.e. “patients-zero” or sets of initially infected subjects that will start de epidemy). Dispersion measures as standard deviation indeed reflect the “range of freedom” among all possible evolutions concerning those possible initial conditions. So, these measures equate to confidence intervals in conventional models.

As the network models are realistic and dynamic, the reproductive number of the epidemic does not need to be adjusted in the equations, as in conventional models. Because this model reproduces the dynamics from where reproductive number is changing as more subjects have their outcomes (immune/deceased) and inter-people connectivity is modulated by factors as a lockdown. This is an advantage of this modeling strategy.

Model validity was checked by a comparative analysis of behavior characteristics previously determined for the population or real data about epidemy evolution, and data from patterns (i.e. median or means) obtained with many simulations (as explained above). In other words, are two ways to prove the representativity of the model. First, for theoretical scenario, extracting the coefficient of exponential growth from the simulated incidence curve and verifying if corresponds to adopted values to R0. For *R0* values found in literature (Sanche et al., 2020) in this model (Ma et al, 2013), we obtained incidence curves that fit (best-fit) an exponential with growth rate parameters that are nearly the same as those of the actual incidence curve. Using the exponential model (equation 4), we found *r* 0.16 and 0.28, respectively, for R0 equal to 2.7 and 5.7, inside or near the respective intervals previously estimated for these R0 (Sanche et al, 2020). Therefore, this model might be considered representative of the evolution of an epidemic such as that of COVID-19 a priori. Second, for real scenario, we simply fit empiric data serie, representative to epidemic progression (here, cumulative mortality counting), upon simulated data. We found overlapping curves with small estimated variance, nearly 2.5%. In both scenarios, we can regard this model as representative to COVID-19 epidemics.

### About Theoretical scenarios

Simulations about theoretical scenarios did not estimate objective parameters of lockdown coverage and time. However, due to its validity and realistic nature for homogeneous and isotropic populations, this model might possibly be useful for quantitative estimates in further studies. A qualitative evaluation performed with our model using current parameter for a theoretical population with low and high R0 values indicates some paradigms for lockdown effectiveness: (1) even in ideal lockdown coverage conditions (90%), only periods longer than 5 weeks would be effective to control the epidemic; (2) softer coverages would only be effective if the epidemic has a lower R0, and even so, for relatively longer periods; and (3) for non-immune population, in epidemics with R0 relatively low, lockdowns with strong coverage (here 90%) did not flatten the curve, although they significantly delay the peak or break the epidemy without population immunization (what occurs using R0=1.44, data not shown).

At first glance, our results from theoretical scenarios seem to conflict with the Chinese experience of 5 weeks of lockdown to control the pandemic. The determining factor is undoubtedly physical lockdown with maximum restriction of urban mobility. However, there are other important factors in the control of epidemic spread: the correct and general use of homemade masks, per se, reduces the transmission of respiratory viruses by approximately 95 to 99% (Sunjaya e Jenkins, 2020). The Chinese people have experienced respiratory virus outbreaks for decades and it is part of their daily lives to have this self-care behavior, which certainly maximizes the effect of lockdown.

### About realistic scenario in Rio de Janeiro

However, the above conclusions are only appliable to lockdowns upon non-imunne populations, with epidemics in course, as in Rio de Janeiro scenario, where the behavior of incidence curves was different. Feeding the model with updated parameters such as Re rather than R0 and the current epidemic status from a specific population, we can infer estimates, and differences between model outputs and real outcomes would be merely scalar.

The local authorities have currently calculated the social confinement nearly to 50% in last 30 days (In Loco, 2020). Under this status (Melan et al., 2020; Secretaria de Saúde do Estado do Rio de Janeiro, 2020; In Loco, 2020) we could predict for Rio de Janeiro the incidence peak in approximately 20 days, with a devastating effect due the total number of active (and severe) COVID-19 cases, nearly 7,5 million of people.

We lost a better outcome, since no heavier lockdown in Rio de Janeiro already is done. However, it is still possible to have a different scenario if we increase the lockdown to 65 to 75% for 70 days, according to our model, reducing the impact of the epidemic by 40%. Thus, intensifying social distance could be necessary.

In Brazil, despite the low lockdown coverage (generally below 50% and never above 60%), we have an estimated reproductive number 1.8 or lower for over 80% of national territory (Perone, 2020). We know that SARS-CoV2 is especially sensitive to heat (Le Page, 2020), and we are still in the hotter seasons in Brazil, which is unfavorable for the propagation of respiratory infections such as COVID (Kissler et al, 2020; Sun et al, 2020). Therefore, due to climatic and seasonal characteristics of Brazil, it is possible that R0 is lower than that estimated for China, in wintertime. Hence, lockdown duration might be the determining factor for the epidemic, even with lower coverage. On the other hand, early release of lockdown might trigger uncontrollable growth of the incidence curve.

### Conclusion

Indeed, observing theoretical and real scenarios, we can conclude that the reproductive number is a determining factor of lockdown characteristics, although lockdown duration is the less dependent factor on virus reproductive number; that is why the priority factor to be included in epidemic containment policies is: lockdown must be relatively long.

Therefore, we interpreted any decrease in likelihood of infection as “lockdown coverage”. In fact, for human beings to be successful in this war against the virus, staying at home is the most important factor, yet it is not enough: creating habits to mitigate transmission must be a priority in people’s daily lives during this tragedy. The paradigms predicted by this model might provide guidance in terms of state policies and individual behavior related to these habits.

## Data Availability

The Dataset was submitted to mendeley.com

http://data.mendeley.com

## Notes

### Competing Interest Statement

The authors have declared no competing interest.

### Funding Statement

This work has no funding support

## REFERENCES

Aronson JK, Brassey J, Mahtani KR. “When will it be over?”: An introduction to viral reproduction numbers, R0 and Re. Centre for Evidence-Based Medicine WebSite, Oxford University. April, 14, 2020. https://www.cebm.net/covid-19/when-will-it-be-over-an-introduction-to-viral-reproduction-numbers-r0-and-re/, acessed at April, 24, 2020.

Britton T. Epidemic models on social networks—With inference. Statistica Neerlandica. 2020;1–20. https://doi.org/10.1111/stan.12203

Chen Shen and Yaneer Bar-Yam, Why a 5-week lockdown can stop COVID-19, New England Complex Systems Institute (March 24, 2020). In: https://necsi.edu/why-a-5-week-lockdown-can-stop-covid-19, accessed at April, 26, 2020.

Bulut C, Kato Y. Epidemiology of COVID-19. Turk J Med Sci. 2020 Apr 21;50(SI-1):563–570. doi: 10.3906/sag-2004-172.

Cevik, M., Bamford, C., & Ho, A. (2020). COVID-19 pandemic - A focused review for clinicians. Clinical Microbiology and Infection. doi:10.1016/j.cmi.2020.04.023

Cucinotta D, Vanelli M. WHO Declares COVID-19 a Pandemic. Acta Biomed. 2020 Mar 19;91(1):157–160.

Haykin S. Neural Networks: a comprehensive foundation. Saddle River: Prentice Hall, 1994. 768p.

IBGE. Estado Rio de Janeiro. 2019. Acessed at May, 13, 2020. https://www.ibge.gov.br/cidades-e-estados/rj.html

In Loco. Mapa Brasileiro da COVID-19: Social distancing Index. 2020. At: https://mapabrasileirodacovid.inloco.com.br/pt/. Accessed at May 13th 2020.

Kissler SM, Tedijanto C, Lipsitch M, Grad Y. Social distancing strategies for curbing the COVID-19 epidemic. MedRXiv, 2020. doi: https://doi.org/10.1101/2020.03.22.20041079.

Le Page M. Will heat kill the coronavirus? New Sci. 2020 Feb 22;245(3270):6–7

Ma, J., Dushoff, J., Bolker, B. M., & Earn, D. J. D. (2013). Estimating Initial Epidemic Growth Rates. Bulletin of Mathematical Biology, 76(1), 245–260.

Mellan TA, Hoeltgebaum HH, Mishra S. et al. Estimating COVID-19 cases and reproduction number in Brazil. Imperial College London (08–05-2020), doi: https://doi.org/10.25561/78872.

Ministério da Saúde do Brasil. Painel Coronavirus (descriptive statistics and datasets). 2020. https://covid.saude.gov.br/ accessed at May 28th 2020.

Perone CS. 01/May – COVID-19 Time varying reproduction numbers estimation for Brazil. Covid Analysis Repository At: https://perone.github.io/covid19analysis/brazil_r0.html. Accessed at May, 1st 2020.

Rothan HA, Byrareddy SN. The epidemiology and pathogenesis of coronavirus disease (COVID-19) outbreak. J Autoimmun. 2020; 109: 102433

Sanche S, Lin YT, Xu C, Romero-Severson E, Hengartner N, Ke R. High Contagiousness and Rapid Spread of Severe Acute Respiratory Syndrome Coronavirus 2. Emerg Infect Dis. 2020 Apr 7;26(7). doi: 10.3201/eid2607.200282.

Secretaria de Estado de Saúde do Rio de Janeiro. Boletins diários do Coronavirus. Acessed at May, 13,2020. https://coronavirus.rj.gov.br/boletins/

Sun Z, Thilakavathy K, Kumar SS, He G, Liu SV. Potential Factors Influencing Repeated SARS Outbreaks in China. Int J Environ Res Public Health. 2020 Mar 3;17(5). pii: E1633. doi: 10.3390/ijerph17051633.

Sunjaya, A. P., & Jenkins, C. (2020). Rationale for universal face masks in public against COVID - 19. Respirology. doi:10.1111/resp.13834

Wibbens, Phebo D. and Koo, Wesley and McGahan, Anita M., How Many More Days of Social Distancing Before Community Transmission is Controlled? A Hierarchical Bayesian Model of COVID-19 by Jurisdiction (April 17, 2020). INSEAD Working Paper No. 2020/21/STR. Available at SSRN: https://ssrn.com/abstract=3578529 or http://dx.doi.org/10.2139/ssrn.3578529

Wu JT, Leung K, Leung GM. Nowcasting and forecasting the potential domestic and international spread of the 2019-nCoV outbreak originating in Wuhan, China: a modelling study. Lancet. 2020 Feb 29;395(10225):689–697. doi: 10.1016/S0140-6736(20)30260-9. Epub 2020 Jan 31.

Zhao, S., & Chen, H. (2020). Modeling the epidemic dynamics and control of COVID-19 outbreak in China. Quantitative Biology. doi:10.1007/s40484-020-0199-0

